# Effects of Preanalytical Sample Collection and Handling on Comprehensive Metabolite Measurements in Human Urine Biospecimens

**DOI:** 10.1101/2024.01.24.24301735

**Authors:** John Braisted, Theresa Henderson, John W. Newman, Steven C. Moore, Joshua Sampson, Kathleen McClain, Sharon Ross, David J. Baer, Ewy A. Mathé, Krista A. Zanetti

## Abstract

Epidemiology studies evaluate associations between the metabolome and disease risk. Urine is a common biospecimen used for such studies due to its wide availability and non-invasive collection. Evaluating the robustness of urinary metabolomic profiles under varying preanalytical conditions is thus of interest. Here we evaluate the impact of sample handling conditions on urine metabolome profiles relative to the gold standard condition (no preservative, no refrigeration storage, single freeze thaw). Conditions tested included the use of borate or chlorhexidine preservatives, various storage and freeze/thaw cycles. We demonstrate that sample handling conditions impact metabolite levels, with borate showing the largest impact with 125 of 1,048 altered metabolites (adjusted P < 0.05). When simulating a case-control study with expected inconsistencies in sample handling, we predicted the occurrence of false positive altered metabolites to be low (< 11). Predicted false positives increased substantially (≥63) when cases were simulated to undergo alternate handling. Finally, we demonstrate that sample handling impacts on the urinary metabolome were markedly smaller than those in serum. While changes in urine metabolites incurred by sample handling are generally small, we recommend implementing consistent handling conditions and evaluating robustness of metabolite measurements for those showing significant associations with disease outcomes.

---

Comprehensive metabolomic profiling provides relative abundances of metabolites in various specimen types. Metabolites comprise small molecules <1500 Daltons and include intermediates and products of metabolism involved in numerous biological processes (e.g., cell structure, signaling, transcriptional regulation, etc.). Notably, metabolites also reflect various components of the exposome including food related components, medications, and other factors that affect human health. In the past decade, analytical technologies have enabled measurement of these metabolites in a high throughput manner, such that hundreds to thousands of samples can be analyzed on a timescale appropriate for studies with larger sample sizes. At the same time, the breadth of metabolites being captured has also increased, such as the ability to simultaneously measure a broader amount of lipid species (1). For these reasons, metabolomic profiling, either alone or in combination with other omics data, is being performed in epidemiological, clinical, and translational research.

Over the last decade, there has been an increase in the use of epidemiologic studies using metabolomics to examine the role of metabolism in health and disease and to identify biomarkers (2,3). In 2014, the Consortium of Metabolomics Studies (COMETS) was established to build infrastructure and encourage collaboration among epidemiologists employing metabolomics in studies with an epidemiologic study design (4). COMETS now has over 70 prospective cohorts and hundreds of investigators using metabolomics as a primary investigative tool in their studies. With the growing interest in using metabolomics in these studies, there have been efforts to understand how preanalytical factors affect findings.

Preanalytical factors include the collection, shipment, storage (temperature and duration), and handling (e.g., aliquoting) of biological samples prior to analysis. Although research studies are designed to minimize the impact of preanalytical factors by standardizing methods, sample collection and handling still inevitably introduce variability into the study. In response to the need for examining the impact of preanalytical factors on metabolite stability in serum, which is a widely used biospecimen in epidemiologic studies, we previously examined how handling conditions (clotting and refrigeration time, number/temperature of thaws) affected observed circulating levels of metabolites (5). We determined that if handling of serum samples varied even modestly by case status, that results can be biased and lead to false-positive findings. Thus, for studies analyzing serum, sample handling should be matched by case status to minimize these effects. Furthermore, we observed that sample handling of serum affects levels of metabolites, thus, steps should be taken to diminish effects.

We note that sample handling conditions are specific to the type of biospecimen used in studies, whether it be plasma, serum, urine, cells, or tissues (6). Urine is a desirable medium for evaluating the metabolome as it is non-invasive, easy to collect, and provides a more global representation of metabolism. It is also worth noting that urine collection protocols tend to be more variable than those used for blood (7) and that issues in sample handling procedures account for the majority of preanalytical errors observed (8). With this in mind, we aimed to characterize sample handling effects on urine in a metabolomics analysis. We collected urine samples from 13 study participants, subjected the samples to various handling conditions commonly encountered in practice (borate, chlorhexidine, or no urine preservative, refrigeration time, and number/temperature of thaws) and examined how each condition affected observed circulating levels of over 1,000 metabolites. Similar to the serum study described above, we examined the impact of sample handling conditions on metabolite measurements using three key metrics, absolute percent, normalized difference, and metabolite abundance correlations. We further performed a simulated case-control study to estimate false positive rates of uncovering altered metabolites.

## Methods

### Study Population

The study enrolled 13 participants (6 men, 7 women) from the area surrounding the Beltsville Human Nutrition Research Center, US Department of Agriculture (Beltsville, Maryland) in 2016. The individuals were recruited from a database of interested volunteers maintained by the Center.

Participants completed a health history questionnaire. In addition, height and weight were measured to determine body mass index (BMI). The eligibility of the participant was based on self-reported medical history from the health history questionnaire, age of 20 to 65 years at beginning of study, and BMI between 18.5-35.0 kg/m^2^, see **Table 1**. Exclusion criteria included the presence of cardiovascular disease, kidney disease, liver disease, gout, hyperthyroidism, untreated or unstable hypothyroidism, certain cancers, gastrointestinal disease, pancreatic disease, other metabolic diseases, or malabsorption syndromes, the participant being unable or unwilling to give informed consent or communicate with study staff, and other medical, psychiatric, or behavioral factors that in the judgment of the Principal Investigator may interfere with study participation or the ability to follow the collection protocol. Diagnosis of disease was based on self-reported medical history.

**Table 1.** Cohort description.

| Characteristic | No. of<br>Persons | % |
| --- | --- | --- |
| Age, years |  |  |
| <30 | 4 | 30.8 |
| 30-55 | 4 | 30.8 |
| >55 | 5 | 38.4 |
| Sex |  |  |
| Female | 7 | 53.8 |
| Male | 6 | 46.2 |
| Body Mass Index<sup>a</sup> |  |  |
| 18.5-25 | 4 | 30.8 |
| 25-30 | 7 | 53.8 |
| >30 | 2 | 15.4 |
| Current smoking | 1 | 7.7 |
| Alcohol consumption<sup>b</sup> | 6 | 46.2 |
| Coffee consumption<sup>b</sup> | 8 | 61.5 |
| Current medication use | 10 | 76.9 |
| Nutritional supplement use<sup>c</sup> | 8 | 61.5 |

<sup>a</sup> Weight (kg)/height(m)<sup>2</sup>
<sup>b</sup> Within the last 48 hours.

Eight of the 13 participants enrolled also participated in our previous study that characterized sample handling effects on serum in a metabolomics analysis (5). The study protocol was reviewed and approved by the Medstar Health Research Institute’s institutional review board (Clinicaltrials.gov identification number NCT02697500).

### Sample collection and processing

Mid-stream spot urine collections were obtained for each participant in the mornings into a sterile urine collection cup with a vacutainer port. Participants were not asked to fast. Aliquots were divided into three separate tubes corresponding to the following preservative treatments: 1) no preservative, 2) borate preservative (BD Vacutainer C&S Urine Tubes, borate 2.63 mg/mL, sodium formate 1.65 mg/mL), 3) chlorhexidine preservative (BD Vacutainer Urinalysis Plus, chlorhexidine 0.4%, sodium propionate 94%, ethyl paraben 5.6%). Tubes were centrifuged at 600xg for 5 min. One mL of urine, with no preservative, from each participant was pooled and mixed to be used as a QC sample, which was run repeatedly with other experimentally treated samples during metabolomics profiling (see section Laboratory assays). Ten aliquots of each of the 3 preservative samples (30 samples total per participant) were transferred into 2mL cryotubes to be subjected to various sample handling conditions, namely refrigeration and free-thaw conditions, as outlined in **Figure 1**. Conditions tested included 3 preservative conditions (no preservative, chlorhexidine, and borate), 2 refrigeration conditions (24-hour refrigeration, no refrigeration/snap freezing), and 7 freeze-thaw conditions (no freeze-thaw, thaw on ice once or 4 times, thaw in refrigerator once or four times, and thaw at room temperature once or 4 times).

**Figure 1.**
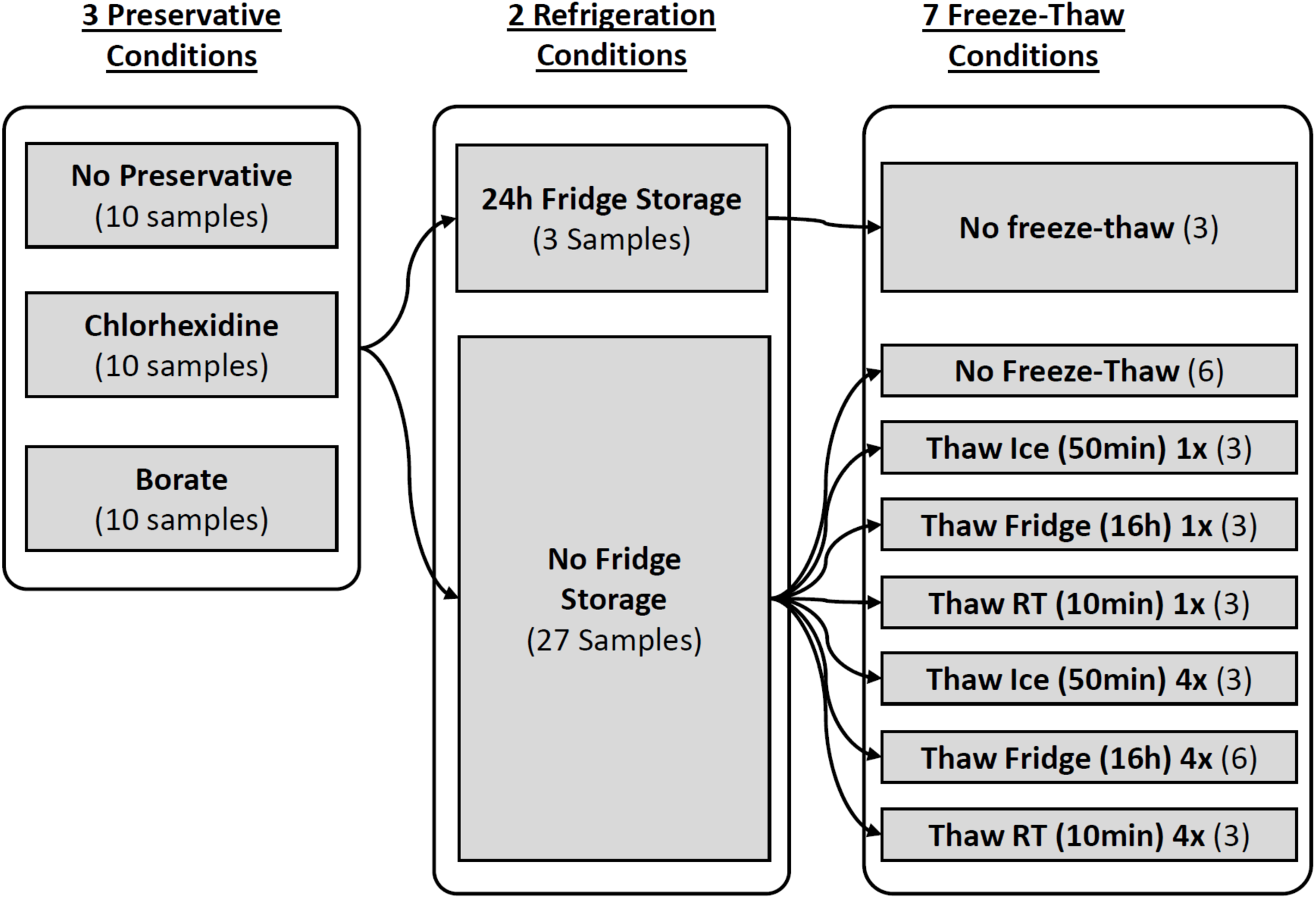
Sample Collection and Preanalytical Sample Handling for Each Participant. Urine samples from each study participant were collected in sterile cups and subsequently divided into 3 preservative condition tubes containing no preservative, borate, or chlorhexidine. Samples were then subjected to various experimental conditions including 24 hours of refrigeration storage, no freeze-thaw, thaw on ice, thaw in refrigerator, or thaw at room temperature for 1 or 4 freeze-thaw cycles. In total, 390 samples (30 samples per participant) were sent for metabolomics profiling.

One sample of each preservative treatment was placed in the refrigerator for 24 h before freezing in liquid nitrogen and stored at -80 °C. Remaining samples were flash frozen in liquid nitrogen and stored at -80 °C. They were then subject to various thawing conditions and freeze-thaw cycles. Thawing was performed at either room temperature, in a refrigerator, or on wet ice. Freeze thaws consisted of either a single thaw or 4 consecutive freeze-thaw cycles. Samples undergoing freeze thaws were snap frozen in liquid nitrogen following their thaw. Samples undergoing freeze-thaw cycles had the thaw time standardized based on previous work (5).

Thaws on ice were fixed at 50 min, room temperature thaws were fixed at 10 min, and refrigerator thaws were fixed at 16 h to simulate a 5 PM to 9 AM thaw often used in laboratories.

### Laboratory assays

In total, 416 frozen samples were shipped to Metabolon, Inc. (Morrisville, NC) for extraction and metabolite profiling. These included 13 participants x 3 preservatives x 10 refrigeration/freeze-thaw conditions + 26 pooled QC samples. Samples were run in batches such that each participant’s 30 samples were run sequentially on the same day, with a pooled QC sample run in positions 3 and 24 of each participant’s sample batch. Pooled QC samples were created from 1 mL aliquots with no preservative from each participant. Urine osmolality was determined at pre-analysis thaw for each sample and reported values as mOsm/kg water. The Metabolon platform uses ultra-high-performance liquid chromatography coupled with mass spectrometry as previously published (9). Metabolon performed the peak picking, alignment, identification, and quality control as previously described (9).

The delivered analytical data matrix consisted of relative abundance levels of 1,208 metabolites, of which 671 were named, as confirmed with a standard, and 32 were described as high confidence identification without a chemical standard. We omitted 160 metabolites that had missing or below detection limit values in 80% or more of 390 study samples, leaving 1,048 metabolites for analysis.

All treatment comparisons were done pairwise within a participant’s set of samples. Missing value imputation was thus performed by participant to eliminate biases introduced from using other individuals’ data for imputation. The method for missing value imputation was dependent on metabolite data coverage over the individual’s 30 samples. For metabolites having sparse data, likely to be missing due to low detection limit (missing in more than 30% of the samples), values were imputed with the half-minimum value for that metabolite in all of the participant’s samples. For metabolites with more complete data coverage (missing in less than 30% of the samples), likely to be missing due to analytical or data preprocessing errors (e.g. co-eluting peaks and challenges in peak picking, etc.), metabolites were imputed using a singular value decomposition approach using the *impute.svd()* method from the R *bcv* package (v. 1.0.1.4)

(10). The median correlation between duplicate gold-standard condition samples was 0.94, similar to prior studies (11, 12). The resulting data matrix was then centered and scaled for unsupervised clustering analysis (PCA) and natural log transformed for downstream statistical analysis. The preprocessing steps are shown in **Supplementary Figure S1**.

### Statistical Analyses

Four previously described and distinct metrics were calculated to quantify the impact of sample handling conditions on metabolite abundances (5). The first metric, the absolute percent difference (APD), reflects the average difference in a metabolite’s log abundance for a particular sample handing condition while keeping all other conditions constant ( e.g. all condition combinations of refrigerator storage and thaw conditions are considered when a particular preservative is evaluated). Second, the normalized difference (ND) estimates the mean difference, in log metabolite level, normalized by between-individual variability. Third, metabolite abundance correlations (Pearson) were calculated for each metabolite to compare a predefined condition (e.g., preservative) against others (e.g., no preservative) while keeping all other conditions constant. The fourth metric estimates false positive rates that result from a simulated case-control study where a given portion of the cases are simulated to have differing sample handling conditions. **Supplemental File 1** provides details on the calculations that result in those metrics.

### Comparison of Sample Handling Effects Across Different Biospecimen Types (Serum and Urine)

APDs from this study in urine were compared to serum stability data from McClain et al. (5) in which the same handling conditions were applied for refrigeration storage and freeze-thaw conditions. Eight of our study participants also participated in the serum analysis. A Wilcoxon Rank Sum test was conducted between the APD values for the 628 metabolites from serum vs. the APD values for the 1048 metabolites from this urine study. This test was run for each of the 7 conditions in common between the two studies, including the 24h refrigeration storage and the six freeze-thaw conditions. The p-values were adjusted using a Bonferroni correction.

## Results

### Data Quality Assessments

Unsupervised clustering of samples, based on 1,048 metabolites measured, confirmed that interindividual variability in metabolite levels dominates the observed variance, not sample handling effects (**Supplementary Figure S2A**). Moreover, participant samples were tightly clustered with no observed outliers, suggesting high quality data.

Missing value assessments demonstrated that sample dilution as indicated by osmolality was a strong determinant of the number of metabolites detected in samples (**Supplementary Figure S2B**. Participants of the study were not fasted, and urine sample osmolality ranged from 132 to 1265 mmol/kg (overall median 440 mmol/kg). The total number of detected metabolites decreased quickly for samples below 500 mmol/kg.

Metabolites missing in >20% samples fell into specific class categories (**Supplementary Table S1**). Specifically, 20% or more metabolites falling under the classes of xenobiotics, cofactors and vitamins, peptides, and lipids were inconsistently represented across samples. Xenobiotics are the compound class with the highest percentage of inconsistently represented metabolites.

### Global Impact of Preanalytical Sample Handling Conditions (APD and ND)

The median and inter-quantile ranges (IQRs) of the absolute percent difference (APD) and normalized difference (ND) for each metabolite provide a summary effect of a treatment across all metabolites. These metrics help discern the most labile metabolites under a given sample handling condition with respect to the gold standard condition (no preservative, no refrigeration storage, no freeze thaw). Median and IQR of APD and ND metrics for samples undergoing various sample handling conditions are shown in **Table 2**. Values are presented for the thaw conditions either with or without preservative treatment. Collectively, the effects of different handling conditions were rather small with median APDs ≤ 5.31 and NDs < 0.05. Also, the median APDs for thaw conditions were generally higher without preservatives. For example, the 4 freeze-thaws on ice median APD increased from 3.79 to 5.31 when computing APD on the subset of samples that lacked preservative treatment (adjusted *P* =1.15E-10, Cohen’s d effect size =0.36). Median and IQR values for ND show a similar trend as observed in APD. It should be noted that gold standard treatment technical replicates, also shown in **Table 2**, have an APD of 3.57 and an ND of 0.039. Most treatments fall close to this APD and ND, suggesting that the treatments are close to or within technical variation when considering the median APD of a treatment.

**Table 2.** Impact of preanalytical sample handling conditions on metabolite abundances.

|  | Absolute Percent Difference (APD) |  |  |  | Normalized Difference (ND) |  |  |  |
| --- | --- | --- | --- | --- | --- | --- | --- | --- |
|  | With Preservative |  | Without Preservative |  | With Preservative |  | Without Preservative |  |
|  | median | IQR | median | IQR | median | IQR | median | IQR |
| **Preservative** |  |  |  |  |  |  |  |  |
| None (Gold Std. Duplicates) | 3.57 | (1.5-7.45) |  |  |  |  |  |  |
| Borate | 2.95 | (1.06-8.60) |  |  | 0.023 | (0.010-0.075) |  |  |
| Chlorhexidine | 3.00 | (1.16-7.31) |  |  | 0.022 | (0.006-0.057) |  |  |
| **Refrigeration** | 2.40 | (0.91-4.92) | 3.01 | (1.22-7.74) | 0.025 | (0.01-0.049) | 0.044 | (0.016-0.088) |
| **Thaw** |  |  |  |  |  |  |  |  |
| 1 thaw |  |  |  |  |  |  |  |  |
| On ice | 1.65 | (0.65-3.56) | 3.13 | (1.29-6.69) | 0.015 | (0.006-0.034) | 0.027 | (0.011-0.056) |
| In refrigerator | 2.61 | (1.11-5.74) | 3.75 | (1.46-8.73) | 0.021 | (0.008-0.040) | 0.032 | (0.014-0.064) |
| At room temp. | 2.13 | (0.89-4.53) | 4.00 | (1.5-9.47) | 0.021 | (0.010-0.044) | 0.034 | (0.012-0.083) |
| 4 thaws |  |  |  |  |  |  |  |  |
| On ice | 3.79 | (1.86-6.41) | 5.31 | (2.53-9.03) | 0.035 | (0.019-0.063) | 0.041 | (0.020-0.074) |
| In refrigerator | 2.39 | (0.94-5.61) | 4.18 | (1.79-8.88) | 0.014 | (0.005-0.033) | 0.021 | (0.008-0.051) |
| At room temp. | 2.41 | (0.96-4.73) | 3.85 | (1.7-8.07) | 0.017 | (0.006-0.040) | 0.025 | (0.011-0.057) |

### Similarity of Metabolite Abundances Between Each Sample Handling Condition and the Gold Standard (Pearson Correlations)

Pearson correlations were calculated to evaluate the similarity between metabolite abundances in a specific sample handling condition compared to the gold standard. Median and IQRs of correlations between each sample handling condition and the gold standard across all metabolites are shown in **Table 3**. For all preservative, refrigeration, and freeze-thaw conditions, the median correlations were ≥0.98, demonstrating a high global concordance in the rankings of metabolite abundances when 100% of samples are switched from one handling condition to another.

**Table 3:** Pearson correlation of relative metabolite abundances for a specific handling condition compared to the “gold standard” condition of no-preservative, no refrigeration storage, and no freeze-thaw cycles.

| Comparison | Median | IQR | 20th Percentile (262) | 15th Percentile (157) | 10th Percentile (105) | 5th Percentile (52) |
| --- | --- | --- | --- | --- | --- | --- |
| **Preservative** |  |  |  |  |  |  |
| Borate | 0.98 | (0.98-0.99) | 0.88 | 0.83 | 0.76 | 0.53 |
| Chlorhexidine | 0.98 | (0.98-0.99) | 0.93 | 0.9 | 0.85 | 0.74 |
| **Refrigeration** | 0.98 | (0.98-0.99) | 0.94 | 0.91 | 0.85 | 0.73 |
| **Thaw** |  |  |  |  |  |  |
| 1 thaw |  |  |  |  |  |  |
| On ice | 0.99 | (0.99-1) | 0.96 | 0.94 | 0.89 | 0.76 |
| In refrigerator | 0.99 | (0.99-0.99) | 0.94 | 0.92 | 0.86 | 0.74 |
| At room temp. | 0.99 | (0.99-1) | 0.94 | 0.92 | 0.85 | 0.73 |
| 4 thaws |  |  |  |  |  |  |
| On ice | 0.99 | (0.99-0.99) | 0.95 | 0.93 | 0.86 | 0.76 |
| In refrigerator | 0.99 | (0.99-1) | 0.94 | 0.92 | 0.87 | 0.74 |
| At room temp. | 0.99 | (0.99-0.99) | 0.95 | 0.93 | 0.87 | 0.73 |

### Expected Number of False Positive Associations When Comparing Sample Handling Conditions to the Gold Standard (Case-Control Study)

We performed a case-control study simulation to predict the expected number of false positive associations due to sample handling conditions rather than sample group differences. This analysis helps clarify our ability to extract meaningful metabolite alterations sought after in epidemiological studies given variations in sample handling conditions which are oftentimes unavoidable. We find that sample handling conditions do not have a strong impact on the expected number of false positive associations predicted for realistic scenarios of different sample handling conditions (1, 5, and 10%) (**Table 4**). In fact, 0 false positive associations were predicted when 1% of the cases had an alternate sample handling condition. When 25% of the cases had alternate use of preservatives, 26 metabolites are expected to be false positives with chlorhexidine use and 45 with borate use. Alterations in refrigeration and thaw conditions were predicted to have < 5 false positive associations. When 100% of cases had alternate sample handling, the number of expected false positive associations were substantially increased (≥63). Again, inconsistencies in use of preservatives, particularly borate, showed the was predicted to have the most impact on false positives (136 for chlorhexidine and 168 for borate).

**Table 4.**
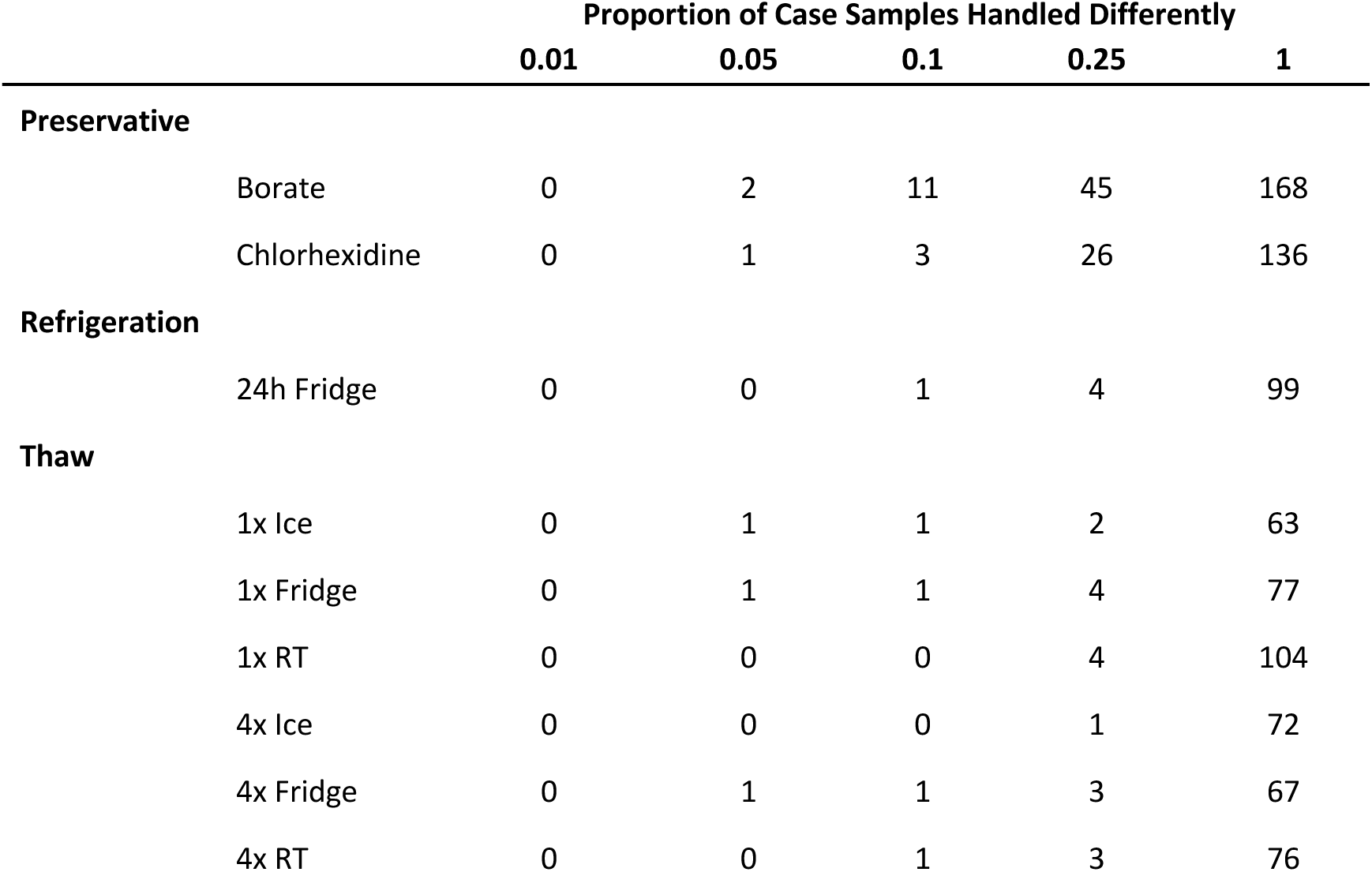
Expected number of false positive associations found when comparing alternate sample handling conditions to the “gold standard” condition.

|  |  | Proportion of Case Samples Handled Differently |  |  |  |  |
| --- | --- | --- | --- | --- | --- | --- |
|  |  | 0.01 | 0.05 | 0.1 | 0.25 | 1 |
| **Preservative** |  |  |  |  |  |  |
|  | Borate | 0 | 2 | 11 | 45 | 168 |
|  | Chlorhexidine | 0 | 1 | 3 | 26 | 136 |
| **Refrigeration** |  |  |  |  |  |  |
|  | 24h Fridge | 0 | 0 | 1 | 4 | 99 |
| **Thaw** |  |  |  |  |  |  |
|  | 1x Ice | 0 | 1 | 1 | 2 | 63 |
|  | 1x Fridge | 0 | 1 | 1 | 4 | 77 |
|  | 1x RT | 0 | 0 | 0 | 4 | 104 |
|  | 4x Ice | 0 | 0 | 0 | 1 | 72 |
|  | 4x Fridge | 0 | 1 | 1 | 3 | 67 |
|  | 4x RT | 0 | 0 | 1 | 3 | 76 |

### Impact of Preanalytical Sample Handling Conditions at the Metabolite Level

While the global impact of sample handling conditions on metabolite intensities in urine is generally low, we found some metabolites are susceptible to changes with handling. The heatmap in **Figure 2** generally shows that the use of preservatives (e.g., borate and chlorhexidine) and four freeze-thaw cycles on ice or refrigerator show the largest differences. Globally, the refrigeration and freeze-thaw cycle conditions resulted in very low numbers of metabolites with significant changes in PD using the established statistical cutoff used above. Further, 4 freeze-thaws tend to increase the number of altered amino acids and lipids for ice and refrigeration pre-treatment. This increase is not observed when evaluating thawing at room temperature or on ice. Compared to the gold-standard samples, borate treatment caused decreased abundance in 5 and increased abundance for 12 of the 28 carbohydrates quantified (based on one-sample t-test, Benjamini-Hochberg (BH) adjusted *P* <0.05). Glucose levels were on average 90% lower with borate treatment. Dehydroascorbic acid is similarly decreased by roughly 90% by borate treatment. Sucrose, lactose and glucuronate were also significantly decreased, consistently over all subjects. Other metabolites that increased with borate treatment included amino acids and derivatives such as glutamine (PD =0.12), glutamate (PD=0.7), cysteine s-sulfate (PD =0.6), 4-hydroxyphenylpyruvate (PD=0.56) and proline (PD =0.26). Chlorhexidine treatment altered PD for 74 metabolites (adjusted *P* <0.05). Nucleotides adenine (PD=-61%) and cytidine (PD=-28%) were decreased with chlorhexidine. **Supplemental File 2** contains statistical results for all treatments, flagging metabolites having significant increased or decreased abundance.

**Figure 2:**
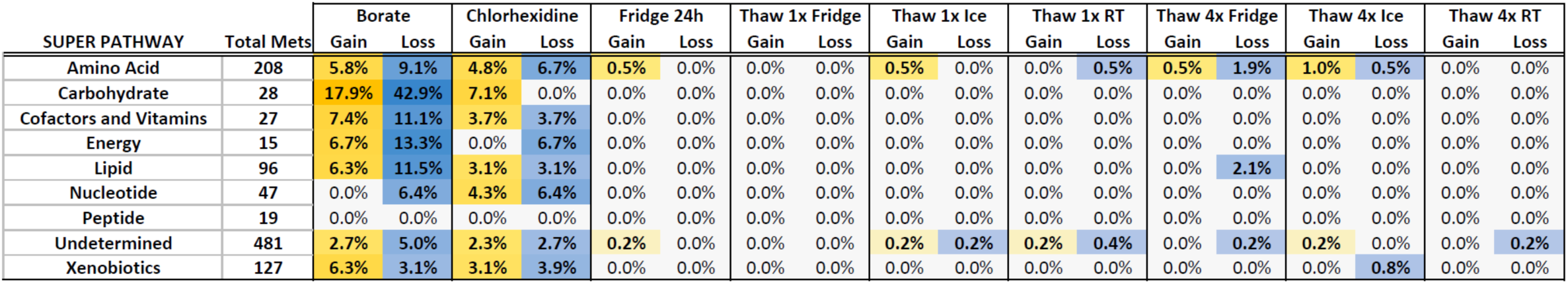
Metabolites and their chemical class altered by preanalytical sample handling. Percentage of altered metabolites within each metabolite class. Metabolites are considered altered if they have a gain or loss of abundance by one-sample t-test (Benjamini-Hochberg adjusted p-value <0.05). Super Pathway annotations are those provided by Metabolon. Gold indicates increases in metabolite abundance while blue elements represent decreased abundance under a given condition. The color intensity indicates the percentage of metabolites having statistically significant abundance changes, with the most intense color indicating that more than 15% of the metabolites in that class have altered abundance, statistically significant percent change.

### Comparison of Pretreatment Effects Between Urine and Serum

When comparing metabolite APD values between urine and those from a previous serum study with identical 24h refrigeration treatment and six freeze-thaw treatments (3), the APD values were higher for serum than for urine for all shared treatments between the studies, except a single thaw performed at room temperature (Wilcoxon Rank Sum, Bonferroni adjusted P < 0.01) (**Supplementary Table S2**). We also note that globally, the number of predicted false positive metabolites in the simulated case-control study were higher in serum samples compared to urine (**Supplementary Table S3**). In the case of 5% of case samples having altered handling, the average number of false positives across the 7 common conditions across the two studies was 24 for serum samples and 0 for urine samples. When the percentage of case samples having altered handling increases to 25%, the average false positive count for serum was 187 compared to urine at 3 false positives.

## Discussion

Results from this study highlight two key findings: 1) urine is strikingly less susceptible to sample handling effects than serum; and 2) preservatives have the largest impact compared to refrigeration and freeze-thaw conditions. Our first key finding is strongly supported by the fact that we followed a study design and analysis scheme used previously for studying serum metabolite stability (5), allowing direct comparison of sample handling effects on metabolite levels in both urine and serum. For the 24h refrigeration and 6 freeze-thaw conditions that were common between the studies, the statistical comparison of serum APD values to urine APD values showed that 6 of the 7 conditions had higher APD values for urine samples. Beyond this, the simulated case-control study had few false positive findings for urine compared to serum. Notably, this finding has practical implications given that many epidemiological studies have collected or plan to collect urine biospecimens. Furthermore, large initatives like the *All of Us* Research Program collect data and biospecimens, including urine, to study many different diseases and conditions. Importantly, while serum is collected more commonly than urine, urine does show important advantages. In addition to facile sample collection and, as we demonstrate here, decreased sample handling effects, metabolites measured in urine are impactful for biomarker detection and for improved detection of diet-related metabolites (13).

The second key finding that preservatives show largest differences in metabolite levels compared to other sample handling conditions is in line with previous observations that while preservatives do prevent bacterial growth, they do not avoid metabolite instability in and of themselves (14). Metabolites commonly evaluated for their effect on human health are impacted by these treatment conditions. For example, dehydroascorbic acid (oxidized form of vitamin C), glucose, glucuronate and glutamine are altered by borate. Similarly, nucleotides are impacted by chlorohexidine.

Further, our results clearly show that consistency in sample handling is more important in reducing false positives than using ideal pretreatment conditions the majority (but not 100%) of the time. In fact, we found that ∼4-15% of urinary metabolites had predicted false positive alterations when 25-100% cases had inconsistent handling conditions. Metabolites most prone to false positives are carbohydrates while those less prone are xenobiotics and amino acids.

For more realistic scenarios (1,5, and 10% differeing sample handling conditions), we found that sample handling conditions did not have a strong impact on the expected number of false positive associations. We do recognize that in practice, consistency in sample handling when thousands of samples are being evaluated is extremely difficult to achieve. Nonetheless, this study highlights the importance of carefully understanding the preanalytical handling conditions to ensure that cases and controls have similar and consistent sample handling.

It is also important to note that all samples for each participant were run as a batch of consecutive injections on the mass spectrometer to minimize the introduction of batch differences between samples within a participant’s sample set. All treatment effects were analyzed within an individual’s sample set, in a pairwise manner to eliminate between-individual differences. This approach of reporting statistics within an individual’s measurements ensured that sample treatment was the main variable being evaluated.

Other studies have evaluated pretreatment effects on the urine metabolome (8, 6, 15). Most studies, consistent with ours, report effects on the urinary metabolome due to refrigeration or freeze-thaw cycles (16, 14) or preservatives (17). However, others report no effects on the urine metabolites due to freeze thaws. One such study was conducted in 6 females and evaluated 63 metabolites (16) and it is thus possible that results are specific to females and to the scope of the metabolome being measured. Another study evaluated the stability of adenosine in a case-control cohort of 40 diabetic patients and 40 healthy controls (18). While limited to a single metabolite, this study is a good example of the importance of evaluating the stability of potential biomarkers derived from epidemiological studies, since, as we observe as well, not all metabolites are affected similarly by pre-analytical conditions. Lastly, when evaluating effects of diet on the metabolome, one study reported that urine metabolites are more prone to variations than blood metabolites (19). While this observation contradicts our global finding that urine metabolites are more robust than blood metabolites in terms of preanalytical differences, we do report that xenobiotics show the highest percentage of inconsistently represented metabolites. This observation could reflect participant differences in diet, medications, or other exposures.

Despite our study approach and design, some limitations are worth noting. First, we recognize that urine samples can be collected through different procedures, from spot to 24-hour urine collection (20). Our study evaluated spot-collected samples, which is most common in epidemiology studies and a less resource-intensive collection method compared to 24-hour urine collections. Notably, our spot urine collections were performed at a similar time of day for each participant, reducing potential biases incurred due to circadian variation (21). Second, our sample size was relatively small and abundance levels between individuals could be affected by osmolality and could therefore impact our ability to observe preanalytical effects. Low osmolality samples tended to have higher APD than higher osmolality across all conditions (data not shown). While the lower salt (and other osmolyte) levels are not likely increasing the variance, the measurement error is likely to increase as concentrations go down, therefore causing an increase in variance. Third, this paper does not address analytical errors (sample preparations and run batches), although study design considerations and inclusion of appropriate QCs can mitigate those errors. In this study, we largely assumed that these analytical errors were consistent across samples. Lastly, all our samples are drawn from healthy volunteers. However, it is likely that metabolite instability due to preanalytical conditions could be different between diseased and healthy individuals due to sample matrix effects, namely due to different enzymatic activity and microbial contamination (6).

This study confirms the importance of implementing consistent handling conditions in epidemiological studies. While urine is suitable for evaluating the metabolome, and as shown here, is more robust to changes in preanalytical conditions than blood, some metabolites are altered due to preanalytical conditions. Robustness of metabolites that show significant associations with disease outcomes should thus be evaluated.

## Data Availability

Metabolomic data will be made publicly available after acceptance in peer reviewed journal.

## Acknowledgements

This work was supported in part by the Intramural Research Program of the National Center for Advancing Translational Sciences (ZIC TR000410-05), National Institutes of Health.

## Conflict of interest

none declared.

---

**Supplementary Figure S1.**
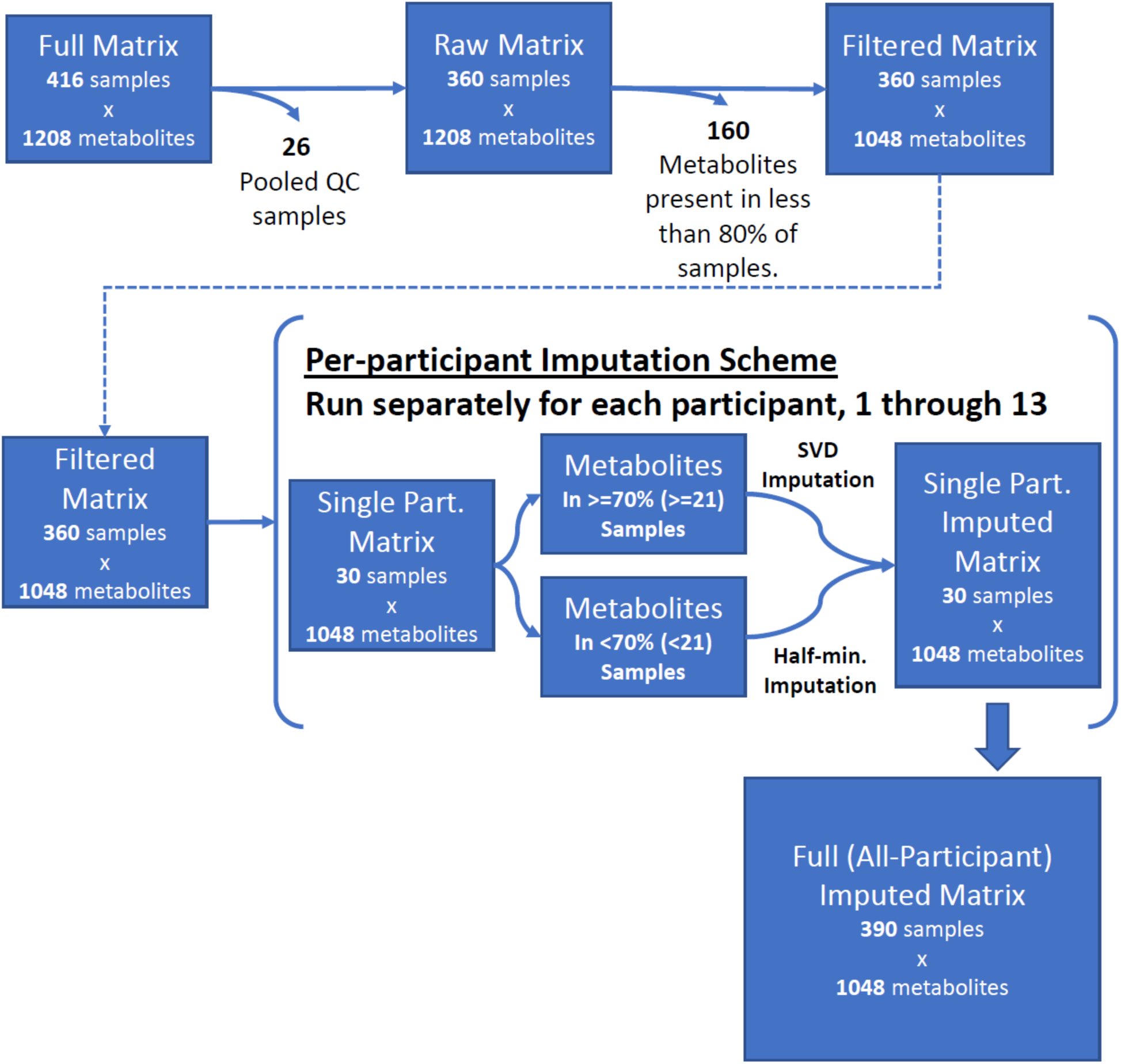
Workflow delineating the data preprocessing steps.

**Supplementary Figure S2:**
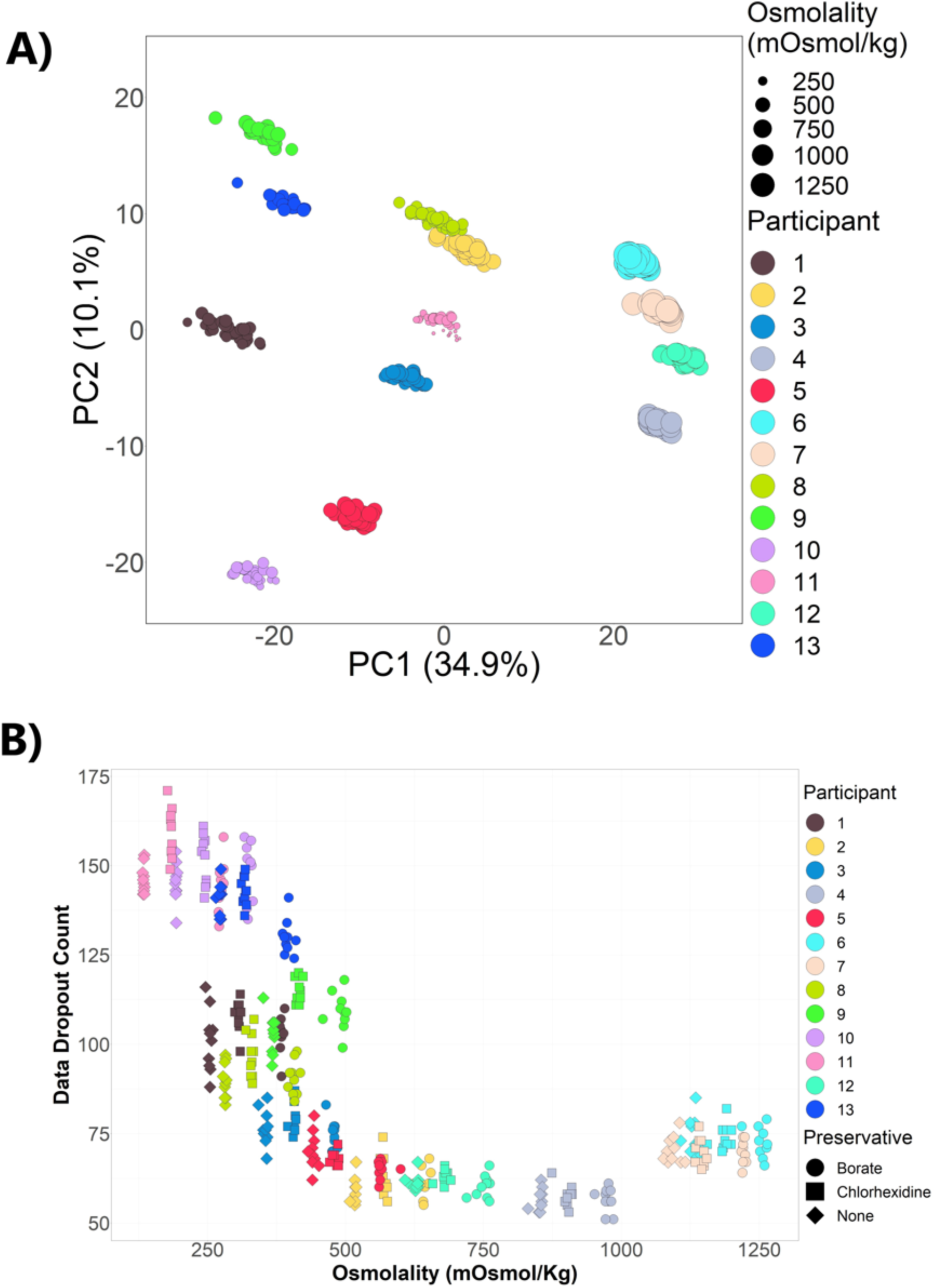
Data quality assessment. A. Principal Components Analysis of our cohort metabolite abundance profiles. The input matrix included the log normalized abundances for 1,024 metabolites for each of the 390 samples (excluding QC samples). Colors are associated with participant ID. Point size relates to sample osmolality. B. Number of metabolite missing values for each sample as a function of sample osmolality. Preservative type is indicated by marker shape.

**Supplementary Table S1.** Total metabolites measured and those filtered out due to being missing in more than 80% of 390 samples. Super pathways are those provided by Metabolon.

| Super Pathway | Total | Missing | Percent |
| --- | --- | --- | --- |
| No Annotation (X-mets) | 537 | 56 | 10% |
| Xenobiotic | 177 | 50 | 28% |
| Lipid | 120 | 24 | 20% |
| Amino Acid | 218 | 10 | 5% |
| Cofactors and Vitamins | 35 | 8 | 23% |
| Nucleotide | 52 | 5 | 10% |
| Peptide | 24 | 5 | 21% |
| Carbohydrate | 30 | 2 | 7% |
| Energy | 15 | 0 | 0% |
|  | 1208 | 160 |  |

**Supplementary Table S2.** Wilcoxon Rank Sum statistic comparing metabolite APD values between serum and urine metabolites for conditions common to both studies.

| Treatment | Serum Mean | Urine Mean | Serum Median | Urine Median | pVal | Bonferroni-Adj pVal |
| --- | --- | --- | --- | --- | --- | --- |
| No Refrigeration vs.24 hr Refrigeration | 10.48 | 5.37 | 4.75 | 3.01 | 1.40E-10 | 9.82E-10 |
| No thaw vs 1x thaw on ice | 7.23 | 5.09 | 4.74 | 3.13 | 1.26E-09 | 8.81E-09 |
| No thaw vs 1x thaw in refrigerator | 9.84 | 6.09 | 6.04 | 3.75 | 1.86E-12 | 1.30E-11 |
| No thaw vs.1x room temperature thaw | 7.37 | 6.59 | 4.65 | 4.00 | 1.21E-01 | 8.46E-01 |
| No thaw vs 4x thaw on ice | 13.52 | 6.68 | 10.05 | 5.31 | 2.35E-36 | 1.64E-35 |
| No thaw vs 4x thaw in refrigerator | 15.56 | 6.50 | 7.25 | 4.18 | 9.88E-19 | 6.91E-18 |
| No thaw vs 4x thaw in room temperature | 7.23 | 5.75 | 5.54 | 3.85 | 5.64E-10 | 3.95E-09 |

**Supplementary Table S3.** Number of false positive metabolites from the study simulation comparing serum to urine, for three levels of fraction of case samples having altered handling conditions, 0.05, 0.25 and 1.0.

|  | Proportion of Case Samples Handled Differently |  |  |  |  |  |
| --- | --- | --- | --- | --- | --- | --- |
|  | Serum Study |  |  | Urine Study |  |  |
|  | 0.05 | 0.25 | 1 | 0.05 | 0.25 | 1 |
| No Refrigeration vs.24 hr Refrigeration | 17 | 86 | 217 | 0 | 4 | 99 |
| No thaw vs 1x thaw on ice | 12 | 181 | 447 | 1 | 2 | 63 |
| No thaw vs 1x thaw in refrigerator | 22 | 205 | 459 | 1 | 4 | 77 |
| No thaw vs.1x room temperature thaw | 12 | 170 | 375 | 0 | 4 | 104 |
| No thaw vs 4x thaw on ice | 54 | 381 | 688 | 0 | 1 | 72 |
| No thaw vs 4x thaw in refrigerator | 41 | 196 | 386 | 1 | 3 | 67 |
| No thaw vs 4x thaw in room temperature | 8 | 92 | 244 | 0 | 3 | 76 |
| Average Number of False Positives | 24 | 187 | 402 | 0 | 3 | 80 |

## Notes

### Competing Interest Statement

The authors have declared no competing interest.

### Funding Statement

This work was supported by the National Cancer Institute Division of Cancer Control and Population Sciences and the Intramural Research Program of the National Center for Advancing Translational Sciences (ZIC TR000410-05) of the National Institutes of Health and the United States Department of Agriculture Agriculture Research Service (projects 2032-51530-025-00D and 8040-51530-011-00D). The United States Department of Agriculture is an equal opportunity employer and provider. 
The content is solely the responsibility of the authors and does not necessarily represent the official views of the NIH or USDA.

### Author Declarations

The study protocol was reviewed and approved by the Medstar Health Research Institute institutional review board (Clinicaltrials.gov identification number NCT02697500).

